# Testing out of quarantine

**DOI:** 10.1101/2021.01.29.21250764

**Authors:** Lucy D’Agostino McGowan, Elizabeth C. Lee, Kyra H. Grantz, Lauren M. Kucirka, Emily S. Gurley, Justin Lessler

## Abstract

Since SARS-CoV-2 emerged, a 14-day quarantine has been recommended based on COVID-19”s incubation period. Using an RT-PCR or rapid antigen test to “test out” of quarantine is a frequently proposed strategy to shorten duration without increasing risk. We calculated the probability that infected individuals test negative for SARS-CoV-2 on a particular day post-infection and remain symptom free for some period of time. We estimate that an infected individual has a 20.1% chance (95% CI 9.8-32.6) of testing RT-PCR negative on day five post-infection and remaining asymptomatic until day seven. We also show that the added information a test provides decreases as we move further from the test date, hence a less sensitive test that returns rapid results is often preferable to a more sensitive test with a delay.

## Background

Quarantine of potentially infected individuals is a time-tested approach for controlling epidemics. Since SARS-CoV-2 emerged, the US Centers for Disease Control and Prevention (CDC), World Health Organization and others have recommended a 14-day quarantine based on COVID-19”s incubation period.

Concerns have been raised that this is overly burdensome and provides limited added benefit versus shorter durations that may increase compliance by reducing disruptions to normal activities (e.g., work, school). Using an RT-PCR or rapid antigen test to “test out” of quarantine is a frequently proposed strategy to shorten duration without increasing risk. The CDC recently issued guidance that a 7-day quarantine with a negative RT-PCR test from a sample collected within 48 hours of exit, or a 10-day quarantine with no test, may sometimes be acceptable.(1) However, RT-PCR tests can miss infections, particularly prior to symptom onset.(2) The goal of our study was to quantify the proportion of infections that escape detection under different testing strategies and varying quarantine durations.

## Methods

We calculated the probability that infected individuals test negative for SARS-CoV-2 on a particular day post-infection and remain symptom free for some period of time. This is equivalent to the probability that an infected individual leaves quarantine given a particular test time and quarantine duration. See supplement for details.

To calculate these probabilities, we developed a biologically plausible model assuming that test sensitivity and symptom onset are independently linked to an unobserved time point after infection where viral load crosses a critical threshold (Figure 1). Available sensitivity data is usually indexed to time of symptom onset or an inferred time of infection.(2–4) Using such data directly tacitly assumes test sensitivity is either fully dependent on, or fully independent of, symptom onset timing. More likely, both depend on infection timing and an underlying process of viral replication, and thus, as our model assumes, are partially correlated. We obtain estimates of sensitivity conditional on the timing of infection and symptom onset using empirically derived distributions of the incubation period(5) and test performance(2).

**Figure 1.**
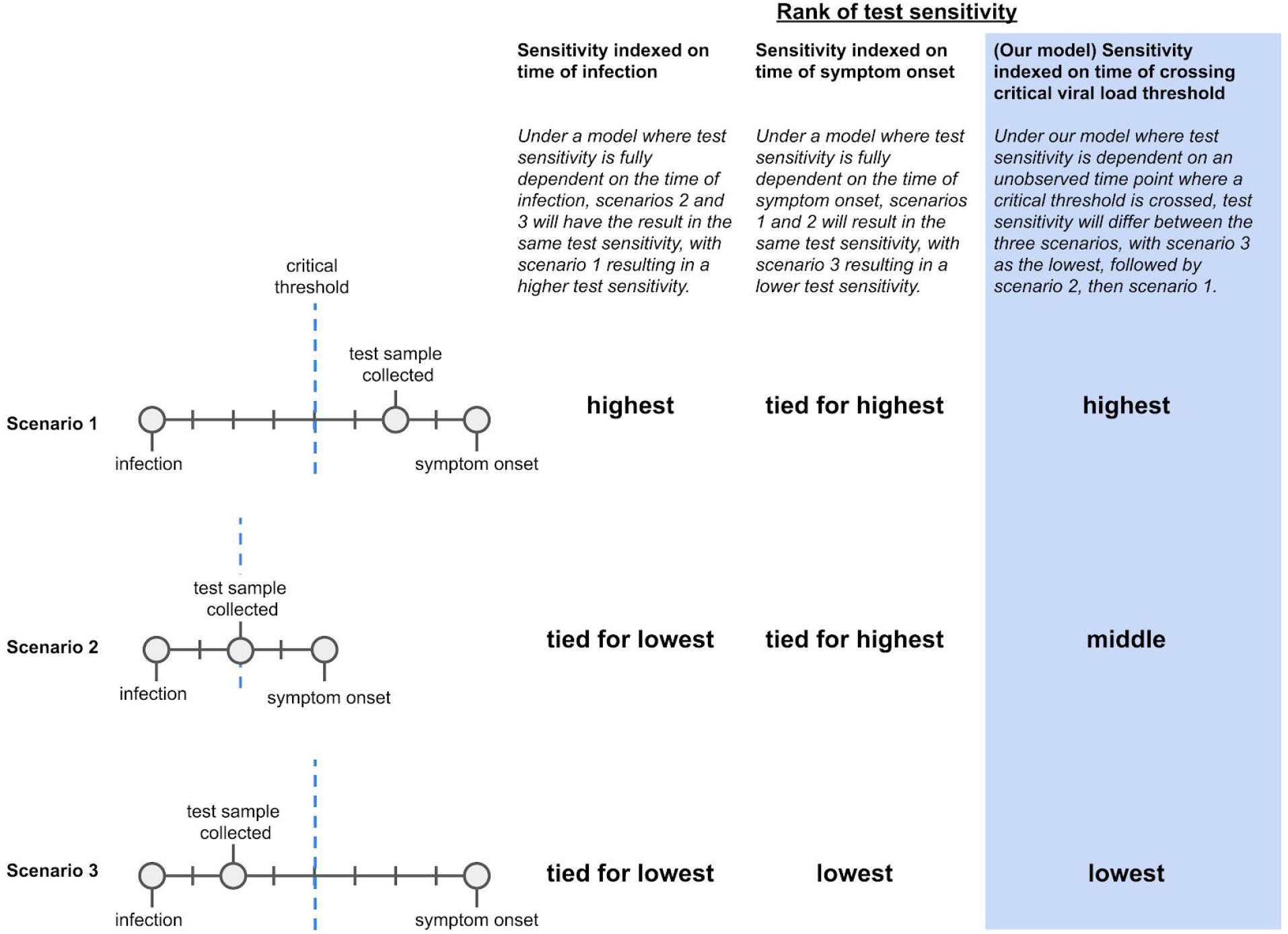
A comparison of our model to other test sensitivity frameworks. Each scenario describes a different relationship between testing and symptom onset, where scenarios 1 and 2 share similar timing between testing and symptom onset, and scenarios 2 and 3 share similar timing between testing and infection time. Only an approach that, like ours, accounts for both dependencies will correctly distinguish between these three cases.

## Results

An RT-PCR test after the first few days of quarantine appreciably decreases the likelihood of an infected individual going undetected (Figure 2A). However, the added information decreases as we move further from the test date, hence a less sensitive test that returns rapid results is often preferable to a more sensitive test with a delay (e.g., Figure 2B). We estimate that an infected individual has a 20.1% chance (95% CI 9.8-32.6) of testing RT-PCR negative on day five post-infection and remaining asymptomatic until day seven, similar to an 8-day quarantine without a test. A negative test on day seven and remaining asymptomatic until day eight misses 9.4% of infections (95% CI 3.1-20.1), similar to a 10-day quarantine, and a negative test and no symptoms on day 10 misses 2.2% (95% CI 0.5-7.4), similar to a 14-day quarantine.

**Figure 2.**
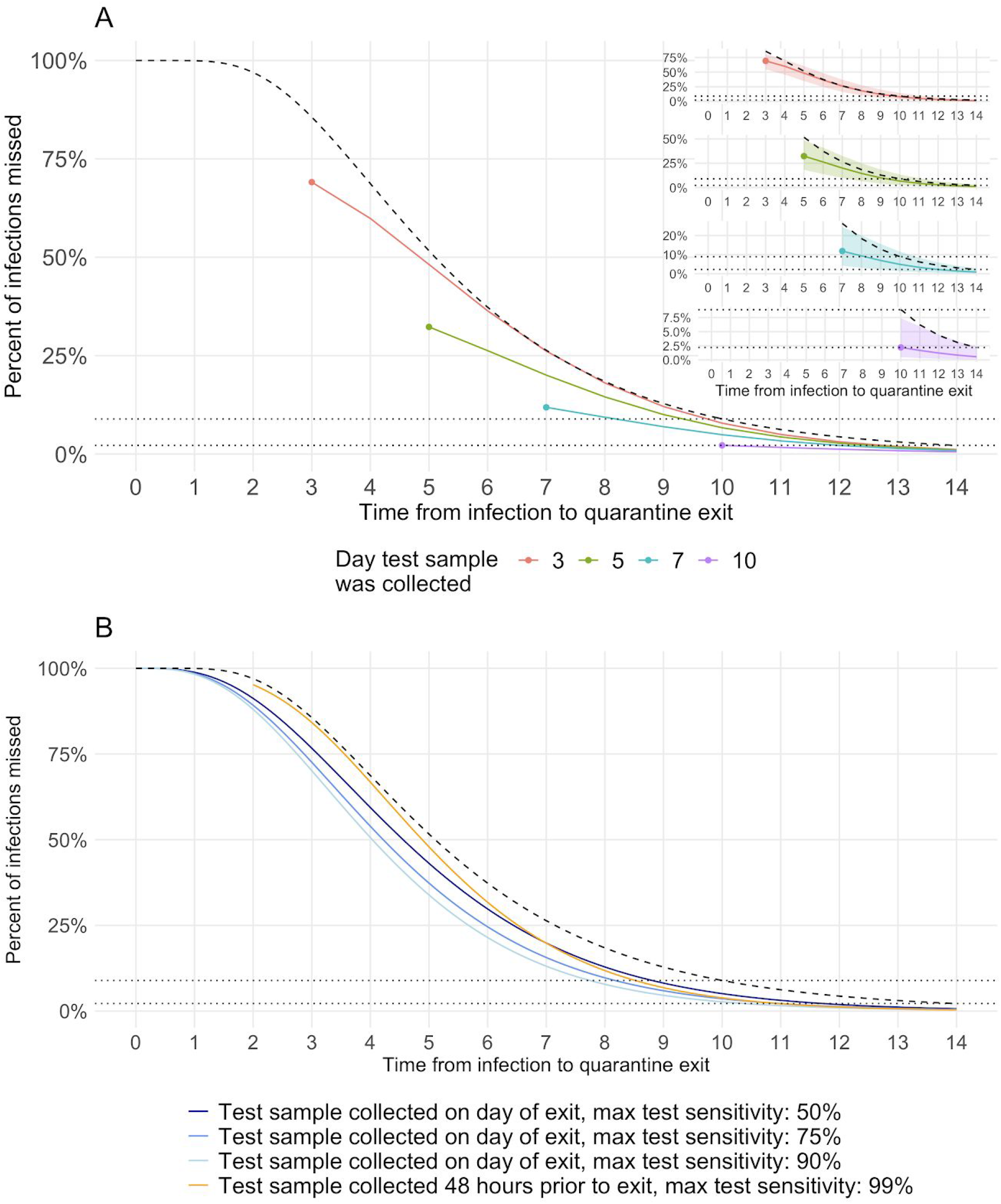
(A) The percent of infections missed is compared between a baseline scenario, quarantining with no symptoms for x days since infection (black dashed line), and 4 quarantine + testing scenarios (solid lines) where individuals without symptoms may exit quarantine when a single negative RT-PCR test sample is collected on either 3 (red), 5 (green), 7 (blue), or 10 (purple) days after infection and remains in quarantine until day x. The circles indicate the day post infection that the test sample was collected. The dotted lines represent a reference point for the proportion of infections missed with a 10 and 14 day quarantine, 8.9% and 2.2%, respectively. (B) Comparison of the percent of infections missed given a test sample was collected 48 hours prior to quarantine exit with a highly sensitive test (orange) to the percent missed given the test sample was collected on the same day as quarantine exit with a less sensitive test (blue).

## Discussion

A negative test can substantially increase the confidence that an individual in quarantine is uninfected, potentially achieving an impact similar to that of quarantines 3-4 days longer. However, the importance of a negative test decreases each day an individual goes without developing symptoms after the test. Hence, a less sensitive test that returns rapid results, and can be taken closer to the end of quarantine, may be more beneficial than a more sensitive test with a delay. The importance of timely test results has been shown to be critical in other contexts(6), and is one reason a negative test on day 5 adds little benefit to 7 days of quarantine without a test.

## Data Availability

Data and R code needed to reproduce the results are provided.

https://lucymcgowan.github.io/covidsens/articles/articles/figures.html

https://github.com/lucymcgowan/covidsens

## References

1. CDC. Options to Reduce Quarantine for Contacts of Persons with SARS-CoV-2 Infection Using Symptom Monitoring and Diagnostic Testing [Internet]. 2020 [cited 2020 Dec 13]. Available from: https://www.cdc.gov/coronavirus/2019-ncov/more/scientific-brief-options-to-reduce-quarantine.html

2. Zhang Z, Bi Q, Fang S, Wei L, Wang X, He J, et al. Insights into the practical effectiveness of RT-PCR testing for SARS-CoV-2 from serologic data, a cohort study [Internet]. Available from: http://dx.doi.org/10.1101/2020.09.01.20182469

3. Wells CR, Townsend JP, Pandey A, Moghadas SM, Krieger G, Singer B, et al. Optimal COVID-19 quarantine and testing strategies. medRxiv. 2020 Nov 30;2020.10.27.20211631.

4. Kucirka LM, Lauer SA, Laeyendecker O. Variation in false-negative rate of reverse transcriptase polymerase chain reaction–based SARS-CoV-2 tests by time since exposure. Ann Intern Med [Internet]. 2020; Available from: https://www.acpjournals.org/doi/abs/10.7326/M20-1495

5. McAloon C, Collins Á, Hunt K, Barber A, Byrne AW, Butler F, et al. Incubation period of COVID-19: a rapid systematic review and meta-analysis of observational research. BMJ Open. 2020 Aug 16;10(8):e039652.

6. Larremore DB, Wilder B, Lester E, Shehata S, Burke JM, Hay JA, et al. Test sensitivity is secondary to frequency and turnaround time for COVID-19 surveillance. medRxiv [Internet]. 2020 Jun 27; Available from: http://dx.doi.org/10.1101/2020.06.22.20136309

