## Supplemental Material for "Testing out of quarantine"

We estimate the proportion of infected individuals who prematurely leave quarantine without being identified as infected due to onset of symptoms or a positive test under a variety of scenarios. Our biologically plausible model indexes test sensitivity based on the time an individual crosses a critical viral load threshold, which is assumed to be related to both time of symptom onset and test sensitivity. To do so, we solve a multi-objective optimization function, simultaneously finding parameters of a gamma distribution that best approximates the incubation period as well as a distribution for test sensitivity. The model fit and an illustrative example are shown in Figure S1.

All calculations were run in R version 4.3.[1] An R package, **covidsens**, is available at https://github.com/lucymcgowan/covidsens as well as code to reproduce the results and figures, available at https://lucymcgowan.github.io/covidsens/articles/articles/figures.html.

*Estimation of proportion of infections missed*

To estimate the proportion of infections missed on a given day post-infection we calculate:

$$\int_{x=0}^{\infty} (1 - s(t-x))(1 - G(t+q-x))f(x)dx$$

Where $x$ is the number of days from infection to crossing the critical viral load threshold; $t$ is the number of days since infection to sample collection; $q$ is the number of days remaining in quarantine after sample collection; $s(t-x)$ is the sensitivity of an RT-PCR test of a sample collected $t-x$ days after crossing the threshold; $g(y)$ is the distribution of days from crossing the critical threshold to symptom onset, $G(y)$ is the cumulative distribution function of days from crossing the threshold to symptom onset; $f(x)$ is the distribution of days from infection to crossing the critical threshold. We examine these estimates across 0 to 14 days post-infection, varying the date the test sample was collected as well as the date of quarantine exit.

*Optimization*

Parameters were determined by first minimizing the negative log likelihood for the incubation period to determine an overall gamma distribution for the incubation period. We then simultaneously minimized the the sum of squared differences between the shape of this incubation distribution and the sum of the shape of the gamma distribution for the time from infection to crossing the critical threshold and the shape of the gamma distribution for the time from crossing the threshold to symptom onset as well as the sum of squared differences between the data on sensitivity[3] and our approximate sensitivity.

*Estimation of sensitivity*

To estimate the test sensitivity we used data from Shenzhen, China collected from 60 symptomatic cases, 57 of whom were diagnosed with COVID-19 by RT-PCR and 3 who later tested seropositive.[3] Among these, 7 had their first RT-PCR test before symptom onset and 20 had their first test on the day of symptom onset. A Bayesian logistic regression model for test sensitivity with a polynomial spline for time since symptom onset was fit to these data (Figure S1B); we then used this fit to find an approximal normalized gamma distribution for the sensitivity indexed based on the day the individual crossed a critical viral load threshold, $s(t-x)$. The sensitivity of the test administered on a particular day after crossing the critical viral load threshold is estimated using a normalized gamma distribution with a shape of 2.25 (SD 0.40) and a rate of 0.21 (SD 0.04), with a fixed maximum of 0.99 (Figure S1E). We then examine how this differs if we vary the maximum sensitivity to 0.5, 0.75, and 0.9 (Figure 2B).

*Estimation of incubation period*

The overall incubation period is estimated to be gamma distributed with a shape of 4.19 and a rate of 0.73. This is comparable to the lognormal distribution suggested in the McAloon et al. meta analysis (Figure S1A).[2] We divide the incubation period into the time from infection until the infected individual crosses a critical viral load threshold, $f(x)$, estimated using a gamma distribution with a shape of 2.09 (SD 0.75) and a rate of 0.73 (SD 0.09) (Figure S1C), and the time from crossing a threshold until symptom onset, $g(y)$, estimated using a gamma distribution with a shape of 2.09 (SD 0.50) and a rate of 0.73 (SD 0.09) (Figure S1D). The estimated distributions $f(x)$ and $g(y)$ were determined through our optimization process, using the empirical sensitivity data[3].

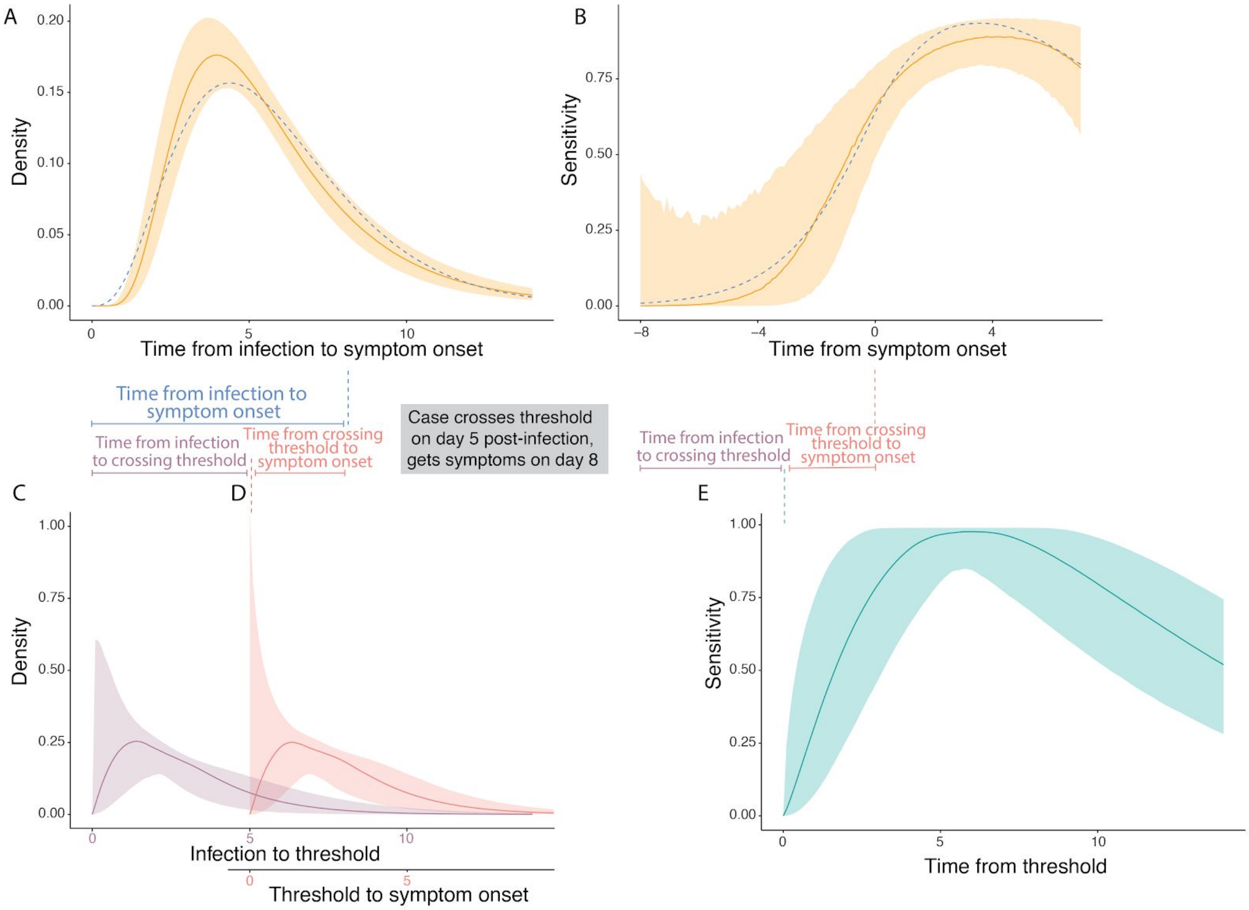


**Figure S1.** Top: (A) Incubation period distribution estimated from observed data (orange) from McAloon et al.[2] and (B) RT-PCR test sensitivity indexed on time from symptom onset (orange) from Zhang et al.[3] compared to model fit (blue, dashed). Bottom: The model fit shown in the top panel (blue, dashed) is generated from three distributions, the combination of which is fit to the above data: (C) the time from infection to crossing a critical viral load threshold (purple), (D) time from crossing the threshold to symptom onset (pink), and (E) RT-PCR test sensitivity indexed by the time since crossing the critical threshold. This bottom panel demonstrates one potential case for illustrative purposes where a case crosses the critical viral load threshold on day 5 and gets symptoms three days later on day 8. The sensitivity curve (E) is indexed on day 5 post-infection (day -3 from on symptom onset), the day they cross the critical threshold. The probability of a missed infection on a given day is then calculated using a combination of these distributions.

### References


1. Team RC. RA language and environment for statistical computing. Versión 3.4. 3, Vienna, Austria, R Foundation for Statistical Computing. Published online 2017.

2. McAloon C, Collins Á, Hunt K, et al. Incubation period of COVID-19: a rapid systematic review and meta-analysis of observational research. *BMJ Open*. 2020;10(8):e039652.

3. Zhang Z, Bi Q, Fang S, et al. Insights into the practical effectiveness of RT-PCR testing for SARS-CoV-2 from serologic data, a cohort study. doi:10.1101/2020.09.01.20182469
